# Describing the landscape of medical education preprints on medRxiv

**DOI:** 10.1101/2023.10.19.23297205

**Authors:** Lauren A. Maggio, Joseph A. Costello, Anthony R. Artino

## Abstract

**Introduction:** A preprint is a version of a research manuscript posted to a preprint server prior to peer review. Preprints enable authors to quickly and openly share research, afford opportunities for expedient feedback, and enable immediate listing of research on grant and promotion applications. In medical education, most journals welcome preprints, suggesting they play a role in the field’s discourse. Yet, little is known about medical education preprints, including author characteristics, use, and ultimate publication status. This study provides an overview of preprints in medical education in an effort to better understand their role in the field’s discourse.

**Methods:** The authors queried medRxiv, a preprint repository, to identify preprints categorized as *Medical Education* and downloaded the related metadata. CrossRef was queried to gather information on preprints later published in journals.

**Results:** Between 2019-2022, 204 preprints were classified in medRxiv as *Medical Education* with most deposited in 2021 (n=76, 37.3%). On average, preprint full-texts were downloaded 1875.2 times, and all were promoted on social media. Preprints were authored, on average, by 5.9 authors. Corresponding authors were based in 41 countries with nearly half (45.6%) in the United States, United Kingdom, and Canada. Almost half (n=101, 49.5%) of preprints became published articles in predominantly peer-reviewed journals. Preprints appeared in 65 peer-reviewed journals with *BMC Medical Education* (n=9, 8.9%) most represented.

**Discussion:** Medical education research is being deposited as preprints, which are promoted, heavily accessed, and subsequently published in peer-reviewed journals, including those specific to medical education. Considering the benefits of preprints and slowness of medical education publishing, it is likely that preprint deposition will increase and preprints will be integrated into the field’s discourse. Based on these findings, we propose next steps to facilitate the responsible and effective creation and use of preprints in medical education.

## Introduction

Peer-reviewed journal articles are the primary form of scholarly communication in medical education. These articles shape medical education discourse, which in turn has implications for educators and administrators who are asked to make critical decisions about what content is addressed in medical school curricula, how learners are taught and assessed, and how educational policy is shaped. However, this form of scholarly communication has been criticized, with researchers highlighting lengthy publication timelines^1^ and the narrow representation of authors.^2-5^ In fields such as physics and mathematics, and increasingly in medicine,^6,7^ preprints have been introduced to mitigate some of these concerns and have become an “integral” component of the scholarly conversation.^8^ Because the majority of medical education journals now allow preprints,^9^ and considering the fact that many other fields and disciplines enthusiastically welcome preprints,^10^ it is important to learn more about this emerging communication mechanism in the context of medical education. Doing so will help to facilitate the responsible creation and use of preprints through informed policies and educational initiatives.

While there is no formal, agreed-upon definition of a preprint, it is generally understood to be a complete version of a research manuscript deposited to a preprint server *prior to peer review* that is immediately and freely available to the public.^11,12^ Preprints have been hailed as a means of empowering authors to: quickly and openly share research, receive expedient feedback, and immediately list research on grant, promotion, and job applications, which can be particularly valuable for early-career researchers.^13^ Preprints are regularly promoted on social media,^14^ reported on by journalists,^15^ and have increasingly become a part of the scientific workflow. For example, a researcher searching Google Scholar would encounter a wide variety of preprints in their results and, more recently, if searching PubMed they will also retrieve preprints reporting research funded by the National Institutes of Health.^16^

Despite the noted benefits, preprint use is not without risks. For example, in clinical medicine, preprints have been criticized for their lack of peer review and for being potentially dangerous to readers who may act on unvetted (and potentially inaccurate and unsafe) claims.^17^ Additionally, from the researcher perspective, there is the risk that a preprint author could run afoul of journal guidelines. For example, if an author initially targets a journal that allows preprints, but their manuscript is rejected, they may find that their work is unsuitable for some journals due to the journals’ incompatible policies.

Recently, Maggio and Fleerackers^13^ pointed out the lack of formal and informal education about preprints in medical education. If preprints are to become a component of our scholarly discourse, then we need to know more about them. Thus, this study asks: what is the current nature of medical education preprints? Our aim is to provide an overview of this emerging communication mechanism with the intent of informing policy and providing opportunities for education.

## Methods

### Data Collection

We gathered medical education preprints and their metadata from medRxiv. medRxiv is “the preprint server for health sciences” — a free, online repository for preprints developed, managed, and owned by Cold Spring Harbor Laboratory, a not-for-profit research and educational institution, Yale University, and the BMJ.^18^ medRxiv accepts preprints from across the health sciences and is organized by categories, which a preprint author selects upon submission. While there are multiple preprint servers, we focused on medRxiv in this study because it is the only server that includes medical education as a category.

We used the R programming package ‘medrxivr’^19^ to establish our foundational dataset of preprints on February 7th, 2023. Using the medRxivr package we accessed medRxiv’s application programming interface (API) to isolate preprints deposited between June 25, 2019 (start of medRxiv) and December 31, 2022, and categorized as *medical education*. As authors can deposit revisions to their preprints, we noted whether or not a manuscript had a revision and, if so, the number of revisions was noted. For our analysis, we focused on the most recent preprint version, which is likely to be the most up-to-date and likely the most complete version. We then downloaded each preprint’s metadata, including: digital object identifier (DOI), title, abstract, publication date, author list, and whether the preprint has been subsequently published. On August 31, 2023, we accessed medRxiv’s API again to check if any additional preprints had been published since our initial query. To enrich our dataset, on March 20, 2023, we manually retrieved the preprint’s altmetrics and article views data from medRxiv.

Next, we queried CrossRef’s API on August 31, 2023 using the R programming package ‘rcrossref’.^20^ From CrossRef, we downloaded the metadata of the published articles that were previously made available as preprints. To provide medical education literature context, we also captured the total number of publications per year for the MEJ-24.^21^ The MEJ-24 is a seed set of journals, which have been described as a core group of journals that represent the field of medical education. To determine the gender of corresponding authors, we submitted their first names to genderize.io,^22^ which is a gender prediction tool. Genderize’s database of names matched 183 of 204 names to a gender with greater than 70% probability. We recognize that these approaches to gender prediction are flawed. First, these approaches present gender as binary, which is an oversimplification of a complex social construct. Moreover, we believe that an individual is the best person to report their gender. However, because self-reported gender data is unavailable, we propose that our approach provides a reasonable method that is in alignment with similar studies.^23-25^ Metadata for these preprints and articles was analyzed using descriptive statistics.

## Results

During the study period, 38,101 preprints with unique DOIs were deposited to medRxiv. Of these, 204 (.5%) preprints with unique DOIs were classified as “Medical Education,” with most preprints deposited in 2021 (n=76, 37.3%) (See Table 1). There were zero revisions for 180 (88.2%) Medical Education preprints and 24 (11.8%) had at least one revision. All preprints were written in English. However, in one instance, a preprint that described an initiative across eight Latin American countries was deposited as a preprint in English, but it was subsequently published as a journal article in English, Spanish, and Portuguese.^26^

**Table 1:** Medical education preprints deposited in medRxiv and articles published in journals between 2019-2023.

|  | 2019* | 2020 | 2021 | 2022 | 2023 |
| --- | --- | --- | --- | --- | --- |
| Preprints deposited | 14 | 60 | 76 | 54 | N/A |
| Preprints published | 3 | 20 | 36 | 37 | 5 |
| Preprints published in MEJ-24 journals (REF) | 1 | 2 | 6 | 8 | 0 |
| Articles published in the MEJ-24**(REF) | 1575 | 3971 | 3585 | 3596 | N/A |

\*partial year - medRxiv began in June 2019.
\*\*excludes *German Medical Science (GMS) Journal of Medical Education* and the *Canadian Medical Education Journal* as they are not indexed in CrossRef

### Preprint Use

On the medRxiv site, users are able to freely access a preprint’s abstract, HTML full-text, and PDF full-text. On average, preprint abstracts were viewed 3,274.42 times (Range 561-128,914; SD=10,311.17) and the HTML full-text 742.45 times (Range 12-96,489; SD=6,803.13). PDF full-texts were downloaded on average 1,875.19 times (Range 63-225,789; SD=15,863.44). The most downloaded preprint was a national survey of medical students in the Philippines to understand barriers to online learning during COVID-19.^27^ This preprint was deposited in July 2020 and has received 236,533 HTML views and PDF downloads. The second and third most downloaded articles were about ChatGPT in relation to the US medical licensure exam (n=120,789)^28^ and the “Reliability, correlations and predictive validity in medical school applicants, undergraduates, and postgraduates in a time of COVID-19” (n=10,183).^29^ In one instance, a preprint examining racial bias in Alpha Omega Alpha inductions was withdrawn upon request of the institution at which the study was conducted due to a lack of ethics review.^30^ While the metadata for the preprint is still available, the preprint’s content is not available for viewing or download. However, there have been over 500 attempts to download the PDF since its retraction. medRxiv provides the public opportunity to post comments on preprints. None of the posted preprints had received comments.

Preprints were featured on social media with an average Altmetric score of 24.09 (Range 1-4,069; SD=285.61) (See Table 2). An Altmetric Score is a numeric score based on social media attention calculated by the Altmetric.^31^ Twitter (now X) was the dominant social media tool used, but preprints were to a limited degree also featured on Facebook, news sites, blogs, video sites, and in policy documents. On average, preprints were tweeted 36.69 times (Range 1-6,384; SD=447.99). The preprint garnering the most altmetric attention was a study describing the performance of ChatGPT on the USMLE,^28^ which was deposited in December of 2022 and covered by 120 news sources and tweeted 6,384 times.

**Table 2:**
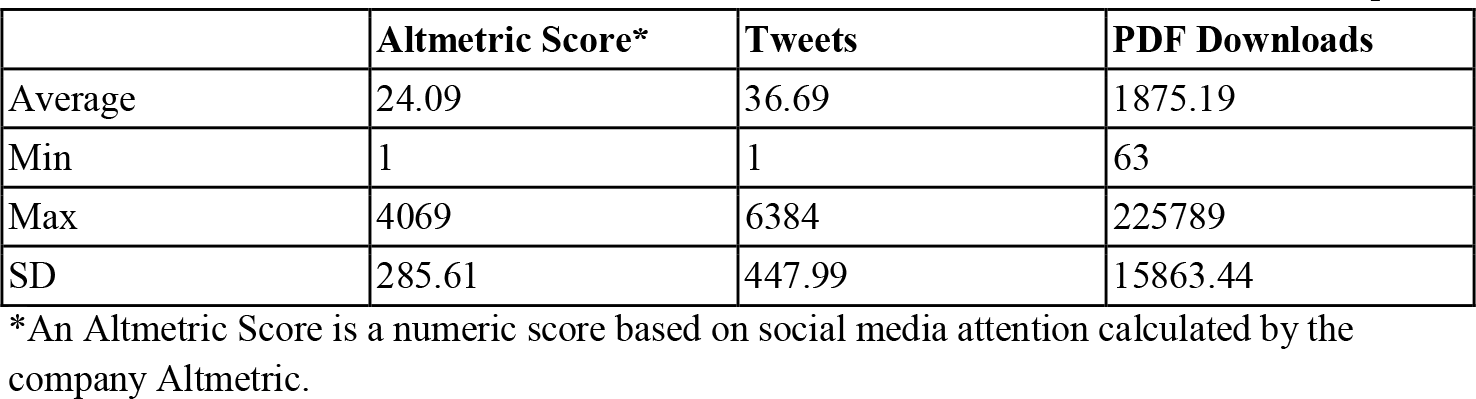
Almetrics, Social media, and Downloads of 204 Medical Education medRxiv Preprints.

### Licensing

Upon depositing a preprint, authors are required to select a copyright license, which signals to potential future users how they may reuse the preprint. medRxiv presents authors with a menu of five copyright licenses specifying the conditions of reuse and one option that does not allow individuals to reuse the content without permission. The majority of authors licensed their preprints (n=147; 72.1%) using a Creative Commons (CC) license. A CC license is a legal tool that provides a standardized way for creators to grant copyright permissions to their work^32^ (for more information on CC licenses, please see Maggio^33^). The most prevalent license selected was the CC BY-NC-ND license, which prohibits commercial use and the creation of derivatives (n=95; 46.6%). Fifty-seven authors did not use a CC license, thereby prohibiting the reuse of their preprint without express permission.

### Authorship

The preprints were authored by 1,220 individual authors of which 1147 were unique authors. Preprints were authored on average by 5.98 (Range 1-34, SD=3.89) authors. The preprint with 34 authors reported on a consortium of 29 medical schools that was assembled to understand the effects of the COVID-19 pandemic on medical schools in the United Kingdom.^34^

Corresponding authors were based at institutions in 41 countries. The United States (n=49, 24.0%), United Kingdom (n=31, 15.2%), and Canada (n=13, 6.4%), were the most represented and included nearly half of the authors. Corresponding authors were predicted to be female (n=83, 40.7%), male (n=100, 49.0%) or undetermined (n=21, 10.3%).

From preprint to published article, 7 articles (7.3%) adjusted author order, but retained the same authorship team. Of these articles, only one changed the first author^35^ and in two cases the last author was shifted to a new author order location.^36,37^ Thirteen preprints (13.5%) had authors added (n=10) or removed (n=3) when they later appeared in a peer-reviewed journal. Of the removed authors, one was ChatGPT, which was attributed for its role in an article about using large language models.^28^

### Publication of preprints

Nearly half (n=101; 49.5%) of the preprints were later published as journal articles, with the most published in 2022 (n=37, 36.6%) and 2021 (n=36, 35.6%). While preprints are immediately available, the average time from preprint to journal publication was 247.04 days (Range 3-770; SD=155.71). Preprints appeared in 68 publications of which 67 were peer-reviewed journals. *BMC Medical Education* (n=9, 8.9%) published the most followed by *BMJ Open* (n=8, 7.9%), and *PLoS One* (n=8, 7.9%). All three journals are peer reviewed. Of the preprints analyzed, eight were ultimately published in MEJ-24 journals accounting for 16.8% of the total published articles. Overall, of the preprints analyzed, 54 were published in journals (79.4%) that are indexed in the Web of Science.

## Discussion

While the current number of medical education preprints is relatively small, this study demonstrates that medical education research is being deposited as preprints, which are heavily accessed and publicized, with many subsequently published in peer-reviewed journals. These findings suggest that preprints likely play a role in our medical education discourse and thus warrant further consideration. We believe that as the benefits of preprints become increasingly known, their volume in medical education will grow, reinforcing the need for medical education researchers and editors to better understand the nature of preprints. In addition, because preprints are only just now emerging in medical education, and are not without risks, we believe there is a need to provide related training and draft informed policies/guidance for their use. In this discussion, we first provide context for our findings and then propose potential future directions.

When compared with the overall volume of publications in medical education,^2^ the current rate of preprints deposited seems quite small. However, preprint usage, as measured by full-text views and PDF downloads, is considerable when compared with the use of published medical education articles. A 2018 study found that, on average, articles published in medical education journals were viewed 881 times as either an HTML page or downloaded PDFs.^38^ Contrast this number with our finding that preprints were downloaded as PDFs alone, on average, 1,875 times and were viewed as HTML pages, on average, 742 times. As access counts such as these are indicators of post-publication impact,^39^ this finding suggests that there is an interest in preprints, which seem to be entering our medical education discourse.

Preprint use may be facilitated by several sources. First, preprints are immediately publicly accessible without any barriers to access (e.g., no passwords required, no need for a reader account). In an international survey of medical educators, the majority of respondents (81%) indicated that having access to medical education literature facilitated the application of research findings to educational practices.^40^ Researchers have shown that paywalls deter readers, even those privileged to have access via institutional subscriptions.^41,42^ Moreover, open access medical education articles have been shown to be more highly accessed than those requiring a subscription.^38^ The publicity of preprints in the media and on social media may also drive their use. Preprints in this study garnered an average Altmetric score of 24.09. Contrast this finding with research showing that published, peer-reviewed medical education articles had an average altmetric score of 7.11.^31^ Part of this difference may be due to the fact that social media attention is directly supported by preprint servers, which, by default, tweet every deposited preprint to a large and broad range of followers across disciplines and fields. This audience is quite different (and more diverse) than that which most medical education journals can reach in their social media feeds. For example, medRxiv’s X account has over 17,000 followers as of September 2023. Additionally, preprints have become integrated into traditional media practices, with many journalists actively seeking and reporting on preprints as part of their normal practice.^15^

Preprints are also adding to the medical education discourse by eventually becoming published articles in a variety of journals, including those specific to medical education (i.e., in the MEJ-24). The practice of preprints subsequently being published in academic journals is not unique to medical education. Reports of this practice from outside of medical education suggest that between 14%^43^ and 42%^44^ of those papers deposited as preprints eventually get published in academic journals. That said, it is unclear how much the preprint changes from being deposited to appearing in a journal, which raises the question of whether or not the findings included in preprints typically change, based on feedback obtained or for other reasons. In other studies, researchers have found that the results often do not change.^45^ Yet, little is known about this practice in medical education, which suggests an area of future research. For example, if researchers were to find that there are minimal changes from preprint to publication in a medical education journal, it raises a question about the value of peer review, which is expensive, requiring huge amounts of dedicated resources from editors and reviewers.

### Future directions

If the number of preprints deposited (and accessed) in the field of medical education continues to increase, then it may be necessary to begin educating our field on their responsible use. First authors, reviewers, and editors must understand what preprints are and how they fit into the field’s scholarly communication processes. They must also be able to identify and critically appraise the potential risks and benefits of preprints to themselves, their journal, and the field. To make such an assessment requires clear policies from journals, funders, and institutions.

Although most medical education journals now include preprint policies in their author guidelines,^9^ policies are less clear at the institutional level. For example, a handful of institutions have begun specifically welcoming the inclusion of preprints in job applications and promotion packages. For example, the University of California, Davis and the New York University School of Medicine both allow preprints to be included as evidence for promotion.^46^ While this is encouraging, we suspect that such policies are rare, leaving it unclear for job applicants and faculty whether or not to list preprints on their curricula vitae. Second, it is important to recognize and consider the responsible use of preprints, which, by definition, are not peer reviewed. Preprint servers play a part in facilitating this awareness. For example, medRxiv preprints feature the warning “Caution: Preprints are preliminary reports of work that have not been certified by peer review. They should not be relied on to guide clinical practice or health-related behavior and should not be reported in news media as established information.”^18^ While this warning is clear, what is less clear is how one should interpret this warning in the context of medical education. For example, should preprints be relied upon to shape educational decision making? Should preprints be an evidence source when no peer-reviewed evidence is available, especially if and when researchers find high concordance between preprints and their eventual peer-reviewed counterpart?^14,45,47^ These and many other questions about preprints and their use should be collectively answered by the medical education community.

In addition to understanding the nature of preprints, it is important that individuals know how to best leverage them. For example, authors have been encouraged to think of a preprint as the “director’s cut” of their manuscript,^48^ which they can share and get feedback on. However, in this study (and, incidentally, in our own practice of preprint posting), we found no evidence of posted comments, suggesting a missed opportunity. While it is very possible that feedback was provided by authors through alternate means (e.g., via social media or direct email to the authors), it appears that we are not yet using this mechanism to the fullest extent. To continue with the comparison of the director’s cut, authors should be aware that preprints do not enforce a word or exhibit limit, unlike many medical education journals. This luxury of space can afford authors the opportunity to provide additional details and data, to include more robust methods sections, additional tables and figures, or more quotes from participants, which can facilitate transparency and encourage study replication.

### Limitations

This study has several important limitations. First, we focused on preprints deposited to medRxiv and did not include alternative repositories, such as bioRxiv or the Open Science Framework, which may contain preprints relevant to medical education. However, our selection of medRxiv, which is the only preprint server focused on biomedicine, enabled us to isolate medical education articles based on their specific classification as designated by their authors. While we attempted to identify all preprints that have been published using CrossRef, we recognize that we may have inadvertently missed some due to the slow speed with which this service ingests records, which can result in delays.^49^ Lastly, we realize that the discourse of any scholarly field is shaped by a variety of sources, with preprints being only one. As such, more research is needed to examine other sources and their influence, such as FOAMed (free open-access medical education).^50^

### Conclusion

The emergence of preprints in medical education may signify a shift in the landscape of scholarly communication. This study reveals the increasing role of preprints in the dissemination of medical education research, highlighting their potential to rapidly share findings and enhance the visibility of research. The considerable access and publicity of preprints, as evidenced by their page views, downloads, and Altmetric scores, suggest they are becoming an important part of our medical education discourse.

However, with the benefits of preprints come some important challenges. The lack of peer review for preprints raises concerns about their content’s credibility. The potential for misinterpretation or misuse of unvetted content underscores the need for critical appraisal skills among readers and the broader scientific community. Furthermore, the complexities surrounding journal policies suggest the need for awareness and education for researchers considering the preprint route.

In summary, preprints represent both an opportunity and a challenge for medical education. As they become more embedded in our scholarly discourse, it is essential that we approach them with a balanced perspective, recognizing their potential to enhance research dissemination while also being aware of the inherent risks. Through informed policies, education, and critical engagement, we believe the medical education community can navigate the ever-evolving landscape of academic publishing, ensuring that preprints serve as a valuable component to our scholarly discourse and scientific advancement.

## Data Availability

All data produced are available online at https://zenodo.org/records/10019574

## Acknowledgments

Reported as none.

## Ethical approvals

This study focuses on publications and does not include human participants. *Disclaimers:* The views expressed in this article are those of the authors and do not necessarily reflect the official policy or position of the Uniformed Services University of the Health Sciences, the Department of Defense, or the U.S. Government.

